# Population-specific polygenic risk for Alzheimer’s disease is associated with Mini-Mental State Examination-based cognitive decline in a Japanese cohort

**DOI:** 10.64898/2026.03.26.26349120

**Authors:** Yutaro Yanagida, Yutaka Nakachi, Issei Morita, Naoto Kajitani, Minoru Takebayashi, Kazuhiro Yoshiura, Manabu Makinodan, Tempei Ikegame, Kiyoto Kasai, Miki Bundo, Kazuya Iwamoto

## Abstract

Alzheimer’s disease (AD) is a major cause of dementia, with polygenic risk scores (PRSs) widely used to capture cumulative genetic risk. While PRSs have been associated with cognitive decline, their relevance to clinically accessible measures in general populations is not yet fully established, particularly in non-European cohorts. In this study, we investigated the association between AD PRSs and cognitive function assessed by the Mini-Mental State Examination (MMSE) in a community-dwelling Japanese older population (*N* = 1,301). Three PRSs were constructed using genome-wide association study (GWAS) summary statistics derived from European and Japanese populations. Among the PRSs, the score based on Japanese GWAS showed the strongest and most consistent association with MMSE score, whereas those based on European GWAS showed weaker or no associations. Stratification analyses further demonstrated that individuals with higher PRS exhibited lower MMSE scores and a higher prevalence of cognitive impairment. Notably, these associations were attenuated after excluding participants with dementia, suggesting that PRS primarily reflects clinically relevant cognitive decline. No significant associations were observed between PRSs and hippocampal volume in our cohort. These findings highlight the importance of population-specific PRS and suggest its potential utility for stratifying cognitive impairment using simple clinical measures in community-based settings.

## Introduction

Dementia significantly reduces individual quality of life (QOL) and social welfare (1,2), with its impact becoming increasingly profound worldwide. Alzheimer’s disease (AD) is the most prevalent type of dementia, accounting for 60% to 80% of all patients (3). Late-onset AD (LOAD) is considered a multifactorial disease, affected by a complicated interplay of genetic and environmental factors (4). Although previous studies have reported the pathogenesis and genetic risk factors of AD including *APOE* gene (5), the overall genetic contribution to onset is not yet completely understood. Notably, genome-wide association studies (GWAS) have identified associations between common single nucleotide polymorphisms (SNPs) and AD (6–9). However, because the effect size of each SNP is very small and a substantial portion of the genetic contribution remains unexplained (10), the polygenic risk score (PRS) has been developed as an individual risk metric that integrates the effects of multiple SNPs identified in GWAS (11).

Beyond its role in capturing polygenic genetic risk, accumulating evidence suggests that PRS is associated with a range of cognitive and neurobiological phenotypes (12–15). A systematic review reported that PRS can distinguish patients with AD from healthy controls and is associated with cognitive impairment, longitudinal decline, and reduced hippocampal volume, although its ability to predict progression from mild cognitive impairment (MCI) to AD remains limited (12). In large cohort studies such as Alzheimer’s Disease Neuroimaging Initiative, higher PRS for AD has been associated with worse baseline memory, accelerated longitudinal decline in memory and executive function, and reduced hippocampal volume (13). Similarly, a UK Biobank study reported that PRS for LOAD was associated with worse cognitive performance, including fluid intelligence, as well as reduced hippocampal volumes (14). While the *APOE* ε4 allele was associated with brain structural changes, it showed no significant association with cognitive performance. Notably, no interaction was observed between the *APOE* genotype and PRS. However, other studies have suggested that PRS-related cognitive decline may interact with age and *APOE* genotype, with effects becoming more pronounced in older individuals, particularly *APOE* ε4 carriers (15). Taken together, these findings support the contribution of PRS to cognitive decline and dementia, as well as its potential utility for risk prediction and stratification in AD.

Previous studies predominantly utilized extensive, multi-domain neuropsychological batteries. However, it remains to be elucidated whether PRS associates with cognitive decline when measured by feasible scales that are routinely implemented in clinical practice. The Mini-Mental State Examination (MMSE) is characterized by its clinical practicality in grading cognitive states (16) and demonstrates robust accuracy for detecting dementia (17). In addition, it is brief, easy to administer, and widely used across diverse clinical settings, facilitating comparability across studies and populations. On the other hand, the MMSE also has notable limitations, including ceiling effects, where high-functioning individuals tend to achieve near-maximum scores, and limited sensitivity to subtle cognitive changes.

Several studies have examined the association between PRS and MMSE, reporting modest associations with cognitive decline (18–20). However, these studies have been conducted predominantly in populations of European ancestry and have largely focused on individuals with dementia or MCI, whereas evidence in general population-based cohorts remains limited. Moreover, the applicability and predictive utility of PRS in Japanese populations remain insufficiently explored. Given potential differences in genetic architecture and environmental backgrounds, validation in diverse populations is warranted.

In this study, we utilized a community-dwelling cohort of older adults from the Arao district, Japan, as part of Japan Prospective Studies Collaboration for Aging and Dementia (JPSC-AD) (21). Using genome-wide SNP data, we constructed three PRSs for AD based on GWAS summary statistics from both Japanese and European populations. Notably, the PRS derived from the Japanese population showed the strongest association with cognitive decline, and best aligned with the distribution of MMSE scores, compared with European-based scores. These findings suggest the importance of population-specific PRS and highlight their potential utility for stratifying clinically accessible cognitive impairment in older adults.

## Materials and methods

### Participants

Participants of the present study were included in the Arao cohort, which consisted of the community-dwelling older people who were 65 years of age or over, living in Arao city, Kumamoto prefecture, Japan (*N* = 1,526). This cohort study is a part of The Japan Prospective Studies Collaboration for Aging and Dementia (JPSC-AD) (21). We excluded participants without consent for genetic analyses (*N* = 5), and analyzed cohort information including age, sex, MMSE score (16,17), and comprehensive SNP genotyping data (**Fig. 1A**). The ethical consideration in this study was approved by ethical review board of human genome and gene analysis in Kumamoto University.

**Fig. 1.**
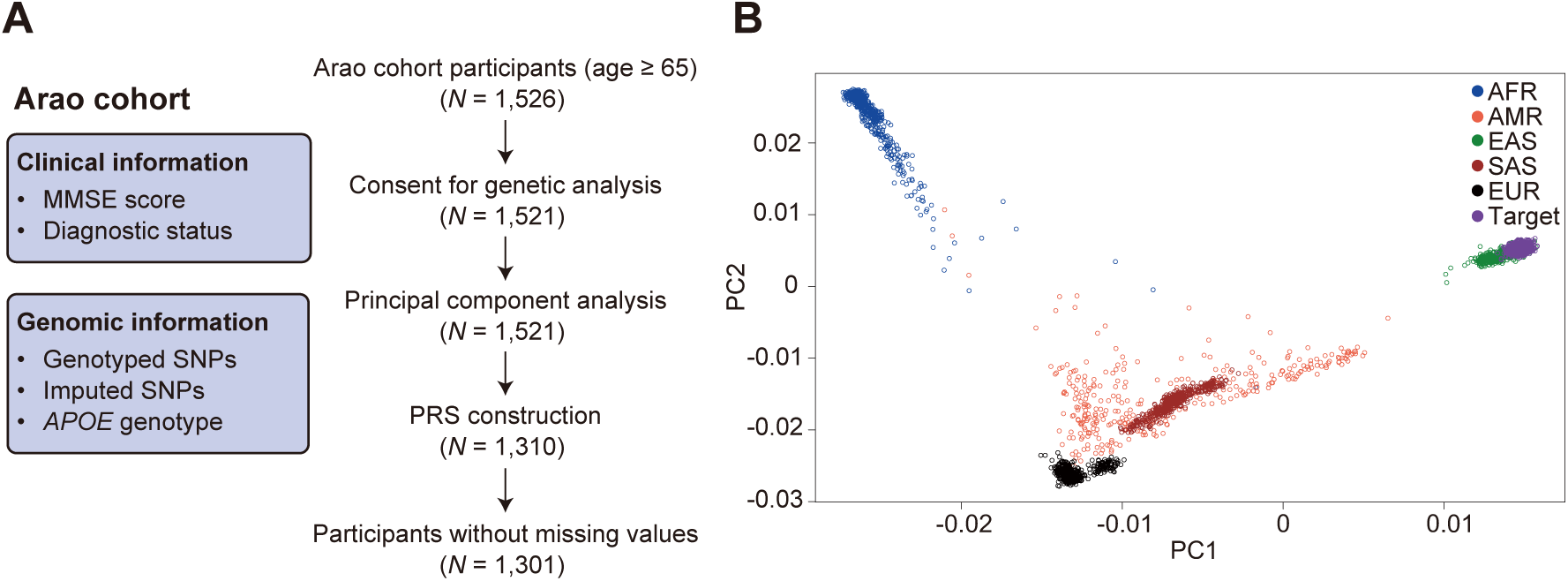
Study design and genetic ancestry of the Arao cohort. **A** Study design and participant selection. Information of the Arao cohort included MMSE score, diagnostic status, genome-wide SNP data, and *APOE* genotype. Of 1,526 baseline participants, 1,521 with consent for genetic analysis were selected and subjected to principal component analysis (PCA). After quality control, 1,310 participants were retained for PRS construction. Participants with complete data were included in subsequent analyses (*N* = 1,301). **B** Scatter plot of the first and second principal components (PCs) for study participants (*N* = 1,521) and the 1000 Genomes project samples (*N* = 2,504). Each point represents an individual based on genome-wide SNP data and is colored by population: AFR, African; AMR, Admixed American; EAS, East Asian; SAS, South Asian; EUR, European.

### Genotyping and imputation of SNPs and *APOE* genotyping

We utilized SNP information genotyped in a previous study (22). In brief, an Illumina Japanese Screening Array (Illumina, San Diego, CA) was used for genotyping of approximately 730,000 SNPs. Quality control (QC) was performed at the sample level and at the SNP level. The SNP imputation was conducted using Minimac4 v1.0.0 (https://github.com/statgen/Minimac4). The reference panels included 1,037 whole-genome sequences from BioBank Japan and 2,504 publicly available whole-genome sequences from the 1000 Genomes Project (Phase 3v5) (23). The *APOE* genotype was identified using a multiplex PCR-based targeted sequencing method, with two SNPs of rs429358 and rs7412 genotyping, as previously reported (24).

### Principal component analysis (PCA) of genome-wide SNPs

We performed PCA using genome-wide SNPs overlapped between the analyzed data in this study (*N* = 1,521) and the 1000 Genomes project Phase 3v5 data (*N* = 2,504) (25,26). We firstly conducted QC as follows: (1) minor allele frequency (MAF) > 0.05, (2) *P*-value of the test for Hardy-Weinberg equilibrium (HWE) > 1.0 × 10^−6^, (3) maximum per-SNP missing < 0.1. We utilized 3,616,282 SNPs for PCA. Data processing for QC and PCA were conducted using PLINK version 1.9 (27) and BCFtools version 1.18 (28).

### Calculation of polygenic risk score

We used three reference GWAS summaries. Two summary statistics from the Psychiatric Genomics Consortium (PGC) were used, one reported in Jansen et al. (hereafter Jans2019) (6), and the other in Wightman et al. (hereafter Wigh202) (8). We also utilized a GWAS summary based on the Japanese population reported in Shigemizu et al. (hereafter Shig2021) (9). QC for SNPs in GWAS summary was conducted as follows: (1) MAF > 0.01, (2) excluding duplicated SNPs, (3) excluding palindromic SNPs. We prepared SNP information of targeted participants in the Arao cohort and utilized overlapped SNPs with reference GWAS summary. In calculation for PRS for AD, we excluded genomic regions flanking of *APOE* genes (hg19 coordinates, chr19:44412079-46412079), following previous studies (15,29), to focus on polygenic effects independent from *APOE*. QC for SNPs in the targeted data was conducted as follows: (1) MAF > 0.01, (2) *P*-value of the test for HWE > 1.0 × 10^−6^, (3) maximum per-SNP missing < 0.01, (4) maximum per-person missing < 0.01 for samples, (5) linkage disequilibrium (LD) pruning, (6) excluding palindromic SNPs. QC for samples was also conducted considering outliers of inbreeding coefficient, relatedness, and sex discrepancy between self-report and estimation by X chromosome. After all, genotyping data of 1,310 participants were utilized for calculation of PRSs. We used MMSE score for phenotype information for calculation of PRS. Covariates for calculation of PRS were age and sex. We constructed PRS using PRSice-2 (30,31) combining each GWAS summary and the target data including SNP genotypes after imputation. Curation of SNP data was conducted using a reference SNP data (GCF_000001405.25.vcf.gz) in Variant Call Format (VCF) downloaded from the National Center for Biotechnology Information (NCBI) dbSNP FTP site (https://ftp.ncbi.nih.gov/snp/latest_release/VCF/) (32,33) and dbSNP 147 summarized data from ANNOVAR (34).

### Neuroimaging analysis

Brain MRI data were acquired using 1.5-Tesla scanners and analyzed for volumetric quantification according to previously published protocols. Detailed information on scanner models, acquisition sequences, imaging parameters, and volumetric procedures is provided in the **Supplementary Methods**.

### Statistical analysis

Analyses were performed in a CentOS Linux release 7.5.1804 environment and by using R version 3.5.1 with in-house scripts (R Foundation for Statistical Computing, Vienna, Austria, https://www.R-project.org/.). The MMSE score was transformed using the natural logarithm, *log(x+1)*, for all correlation and multiple regression analyses. Multiple regression analyses were conducted with standardization. Covariates were age, sex, the first eight principal components (PCs) of genetic background, and the number of *APOE* ε4 alleles. To evaluate the assumptions of multiple regression models, we generated quantile-quantile (Q-Q) plots of the residuals in each model (**Supplementary Fig. S1**). The threshold of statistical significance was set to 0.05.

## Results

### Summary of the analyzed data

We analyzed participants after excluding those without consent for genetic analyses (*N* = 1,521) (**Fig. 1A**). To confirm genetic ancestry, PCA was performed using participant data with the 1000 Genomes Project as reference, showing that all participants clustered within the East Asian group (**Fig. 1B**). We utilized participants without missing values of age, sex, clinical scores, PRS, and the number of *APOE* ε4 alleles in following analyses (*N* = 1,301, **Fig. 1A**). In summary, 501 (38.5%) participants were males, and their ages ranged from 65 to 98 years old, with the mean and standard deviation (SD) of 74.3 ± 6.6. The mean and SD of MMSE score were 27.0 ± 3.2. The distribution of *APOE* genotypes was as follows: ε2/ε2 (*N* = 5, 0.4%), ε2/ε3 (*N* = 109, 8.4%), ε2/ε4 (*N* = 8, 0.6%), ε3/ε3 (*N* = 980, 75.3%), ε3/ε4 (*N* = 185, 14.2%), and ε4/ε4 (*N* = 14, 1.1%).

### PRS associations with MMSE

After QC in PRS construction, we calculated three sets of individual PRSs for AD, based on 904, 96,114, and 208 SNPs, using GWAS summary statistics from Jans2019 (6), Wigh2021 (8), and Shig2021 (9), respectively (**Fig. 2A**). Each PRS was normally distributed (Kolmogorov-Smirnov test, *P*-value > 0.05, **Supplementary Fig. S2**). No significant correlations were observed among the PRSs (*P*-value > 0.05, **Supplementary Fig. S3**). We then evaluated the relationship between PRS and MMSE score (**Fig. 2B**). While no significant correlation was observed between MMSE score and PRS derived from Jans2019 or Wigh2021 (Pearson’s R = 0.04, *P*-value = 0.16 for Jans2019; R = -0.04, *P*-value = 0.17 for Wigh2021), PRS based on Shig2021 showed a significant correlation with MMSE score (R = -0.06, *P*-value = 0.02). We also conducted multiple regression analyses adjusting age, sex, the number of *APOE* ε4 alleles, and the first eight PCs to evaluate influences of the PRS on MMSE score (**Fig. 2C**). Age was significantly negatively associated with MMSE score, whereas female sex was significantly positively associated with MMSE score in all models. *APOE* ε4 allele count showed a significant negative association in the Jans2019 and Shig2021 models and a trend-level negative association in the Wigh2021 model. Among the PRSs, significant negative associations with MMSE score were observed for the Wigh2021 and Shig2021 models, but not for the Jans2019 model. The explained variance of MMSE score, as measured by adjusted R-squared, was 11.1% in the Wigh2021 model, representing a 0.2% increase compared with the covariate-only model, and 11.3% in the Shig2021 model, representing a 0.4% increase compared with the covariate-only model.

**Fig. 2.**
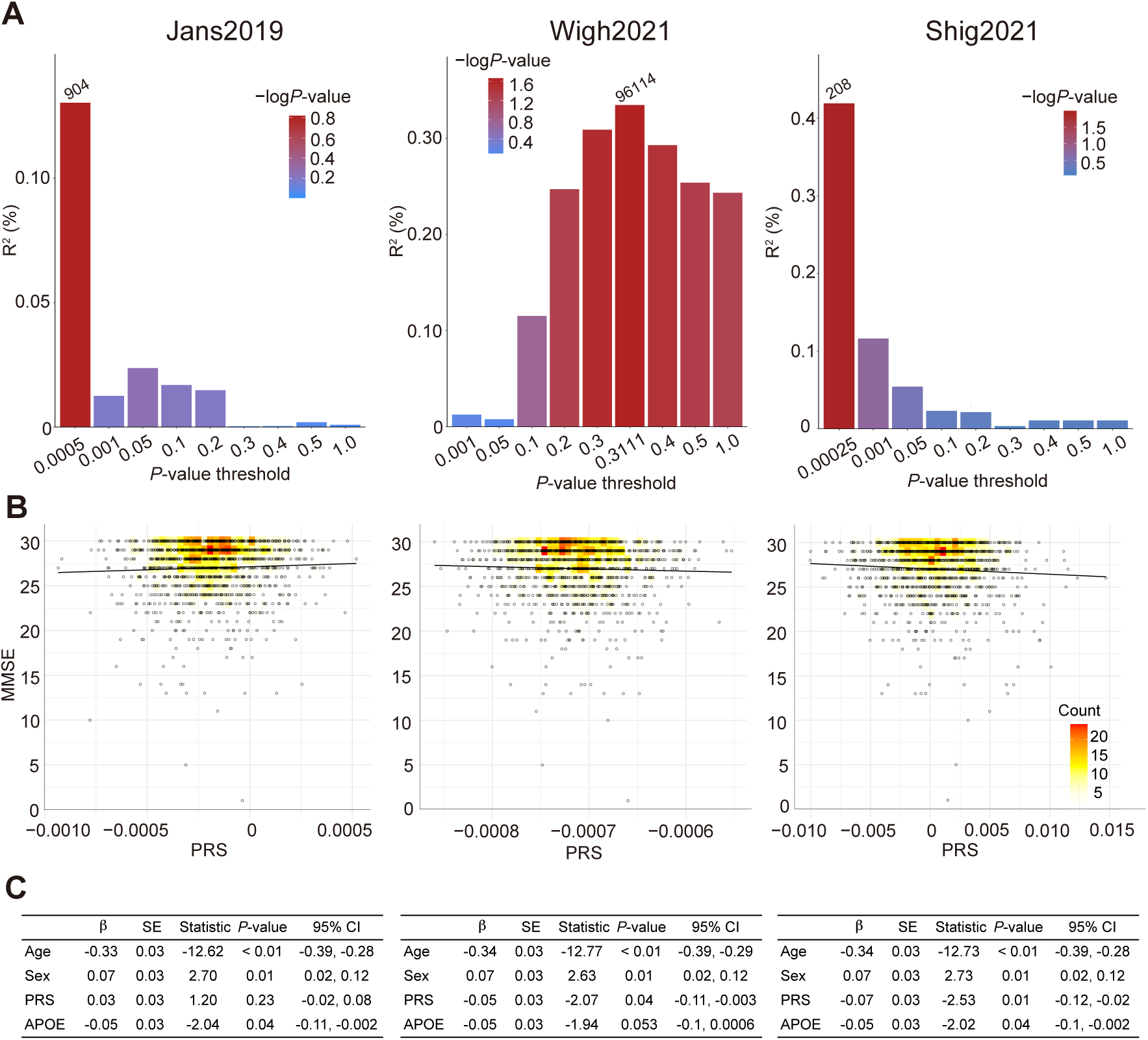
PRS construction and association with MMSE score. PRS construction and evaluation based on GWAS summary statistics from Jans2019 (6), Wigh2021 (8), and Shig2021 (9). **A** Selection of SNPs used for PRS construction across *P*-value thresholds. The horizontal axis indicates *P*-value thresholds applied to GWAS summary statistics, and the vertical axis indicates the coefficient of determination (R^2^, %). Numbers above bars represent the number of SNPs included at each threshold. The color bar represents the -log_10_(*P*-value) of the association with MMSE. **B** Scatter plots of PRS and MMSE score. Color indicates point density, with darker shades of red indicates high density, respectively. Black lines indicate linear regression lines. **C** Standardized multiple regression analysis of MMSE score (*N* = 1,301). Models include age, sex, *APOE* ε4 allele count, the first eight principal components. SE, standard error; Statistic, *t*-statistic; 95% CI, 95% confidence interval.

We additionally performed multiple regression analyses to evaluate the association between PRSs and hippocampal volumes. However, no significant associations were observed (*P*-value > 0.05).

### Stratification by PRS and cognitive decline

The PRS based on Shig2021 showed the strongest association with MMSE score (Fig. 2C) and remained significantly negatively associated after correction for multiple testing (*P*-value = 0.01). We therefore conducted population stratification using this PRS (**Fig. 3A**). We observed a significant difference of MMSE scores between groups defined by PRS deciles (Kruskal-Wallis test, *P*-value = 0.02). Participants in the top 10% of PRS (*N* = 131) showed the lowest median MMSE and significantly or marginally lower MMSE scores compared with most of other groups (Steel’s multiple comparison test). Furthermore, when focusing on the extreme ends of the PRS distribution (top and bottom 1%), a significant difference in MMSE scores was observed between the two groups (Mann-Whitney *U* test, *P*-value = 0.02). In addition, the proportion of participants with an MMSE score of 23 or below, a commonly used threshold for dementia, was higher in the top 1% PRS group (35.7%) than in the bottom 1% group (7.7%), whereas the overall proportion across all participants was 11.6%.

**Fig. 3.**
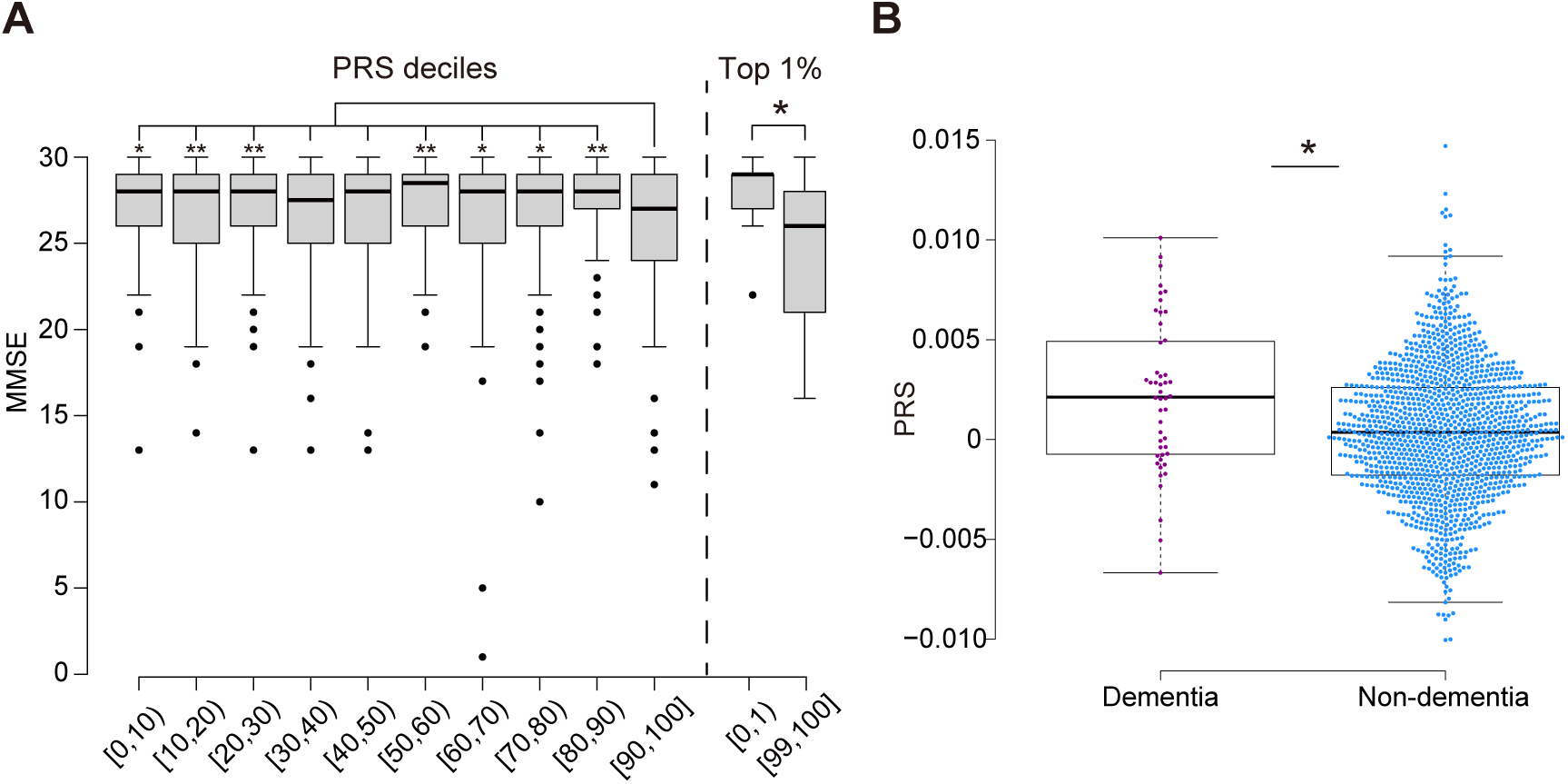
PRS stratification of MMSE and dementia status. **A** Distribution of MMSE scores across PRS percentile groups based on Shig2021 (9), including deciles and the top and bottom 1%. In the decile analysis, asterisks indicate trend or significant differences compared with the top 10% PRS group (Steel’s multiple comparison test; \**P*-value < 0.10, \*\**P*-value < 0.05). In the top 1% analysis, the asterisk indicates a significant difference between the top and bottom 1% groups (Mann–Whitney U test; *P*-value < 0.05). **B** Comparison of PRS based on Shig2021 between patients with dementia (*N* = 48) and participants without dementia (*N* = 1,253). Points represent individual values, and box plots indicate median and interquartile range. The asterisk indicates a significant difference (Mann–Whitney U test; *P*-value < 0.05).

We then focused on participants who were diagnosed with dementia (*N* = 48). These dementia patients showed higher PRSs based on Shig2021 compared to healthy participants (Mann-Whitney *U* test, *P*-value < 0.01, **Fig. 3B**). Analysis excluding these patients left 1,253 participants, and no significant associations were observed between the PRSs and MMSE score in either correlation or multiple regression analyses (*P*-value > 0.05).

## Discussion

To our knowledge, this is the first study to evaluate the association between PRS for AD and cognitive decline measured by MMSE score in a Japanese community-dwelling older population. The PRS based on Japanese GWAS summary statistics showed the strongest association with MMSE, whereas those from European populations showed weaker or no associations. Furthermore, stratification by PRS revealed that individuals with higher genetic risk exhibited lower MMSE scores and a higher prevalence of cognitive impairment.

These findings are consistent with previous studies reporting associations between AD PRS and cognitive decline or neuroimaging phenotypes, mainly in European populations (12–15). However, most prior studies have focused on patients with dementia or MCI and have used extensive neuropsychological batteries. In contrast, our study demonstrates that PRS is associated with cognitive performance even when assessed using MMSE, a simple and widely used clinical tool. This suggests that PRS may capture clinically relevant cognitive differences detectable in routine clinical settings.

A key finding of this study is that the PRS derived from Japanese GWAS (9) outperformed those derived from European GWAS (6,8). This supports the importance of population-specific genetic architecture in PRS construction (35). Differences in allele frequencies, LD structure, and environmental backgrounds between populations may reduce the transferability of PRSs across ancestries. Our results therefore highlight the necessity of developing and validating PRSs within the target population, particularly for clinical applications.

Notably, although the associations between PRS and MMSE were statistically significant, the effect sizes were modest and the increase in explained variance was limited. This is consistent with the polygenic nature of AD, where individual genetic variants contribute only small effects (6–9). Nevertheless, stratification analyses demonstrated that individuals at the extreme ends of PRS distribution showed meaningful differences in MMSE scores and prevalence of cognitive impairment, suggesting that PRS may be informative for risk stratification.

In contrast to some previous reports (13,14), we did not observe significant associations between PRSs and hippocampal volume. Several factors may explain this discrepancy, including differences in sample size, imaging protocols, and population characteristics. It is also possible that PRS effects on brain structure are subtle and require larger or longitudinal datasets to detect.

Importantly, when individuals with dementia were excluded, the association between PRS and MMSE was no longer significant. This suggests that the observed association may be driven primarily by individuals at the more severe end of cognitive decline, rather than reflecting subtle variation within the normal range. This finding indicates that PRS may be more effective in identifying individuals at higher risk of clinically relevant cognitive impairment rather than detecting early or subclinical changes.

This study has several limitations. First, the cross-sectional design precludes conclusions about causality or longitudinal cognitive decline. Second, MMSE has limited sensitivity to subtle cognitive changes and may be affected by ceiling effects. Third, although we adjusted for major covariates including *APOE* genotype and population structure, residual confounding cannot be excluded. Finally, the sample size, particularly for neuroimaging analyses, may have limited statistical power.

Despite these limitations, our findings suggest that population-specific PRS has potential utility as a tool for stratifying cognitive risk in community-dwelling older adults. Future studies incorporating longitudinal data, larger sample sizes, and multi- modal biomarkers will be important to further clarify the clinical applicability of PRS in diverse populations.

## Acknowledgements

We would like to thank the participants for the contribution of their time to the JPSC-AD study. We would like to gratefully acknowledge the diligent work and contributions of all researchers and investigators in the JPSC-AD Study Group.

## Author contributions

YY, MB, YN, and KI contributed to conceptualization. YY, IM, TI, and YN performed methodology development, validation, formal analysis, and investigation. YN, NK, MT, KY, MM, KK, MB, and KI contributed resources. Data curation was conducted by YY, YN, MB, and KI. The original draft was written by YY and KI, and all authors contributed to review and editing. Supervision and project administration were performed by MB, YN, and KI.

## Data availability

The scripts used in the present study are available from corresponding authors on request.

## Supplementary Information

**Supplementary Figure S1.**
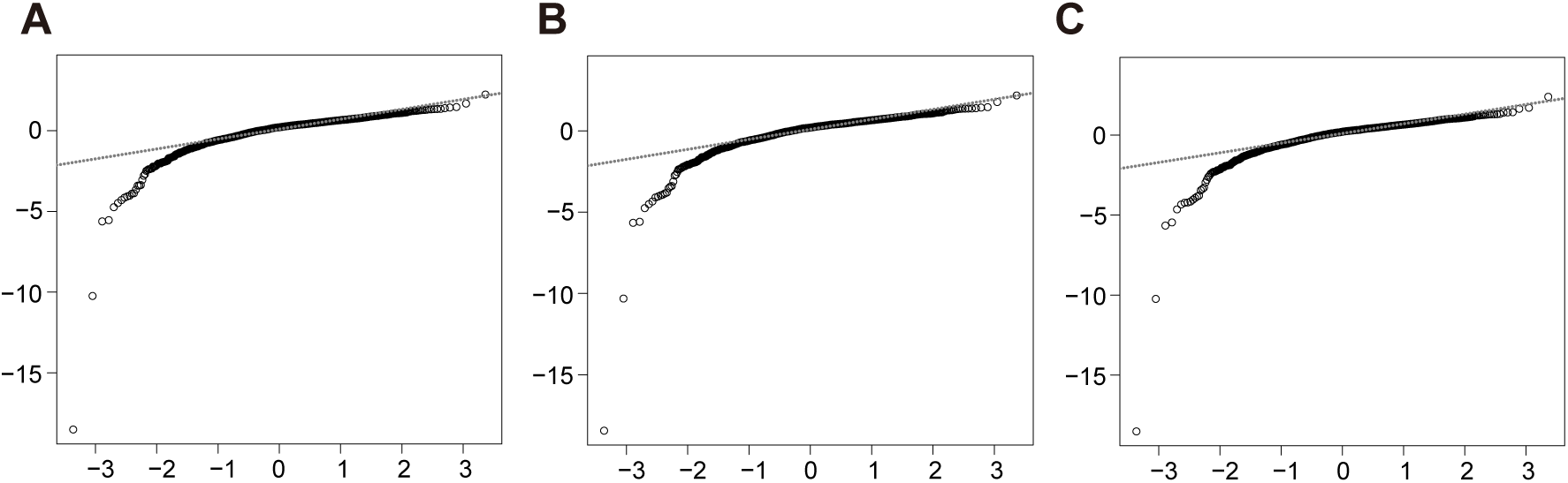
Quantile-quantile (Q-Q) plots of residuals from multiple regression analyses. Panels show Q–Q plots of residuals from regression models of MMSE score. All models included age, sex, the first eight principal components (PCs) of genetic background, and *APOE* ε4 allele count as covariates, with PRSs from Jans2019 (**A**), Wigh2021 (**B**), and Shig2021 (**C**) as explanatory variables. Axes indicate standardized residuals and theoretical quantiles. The dashed line represents y = x.

**Supplementary Figure S2.**
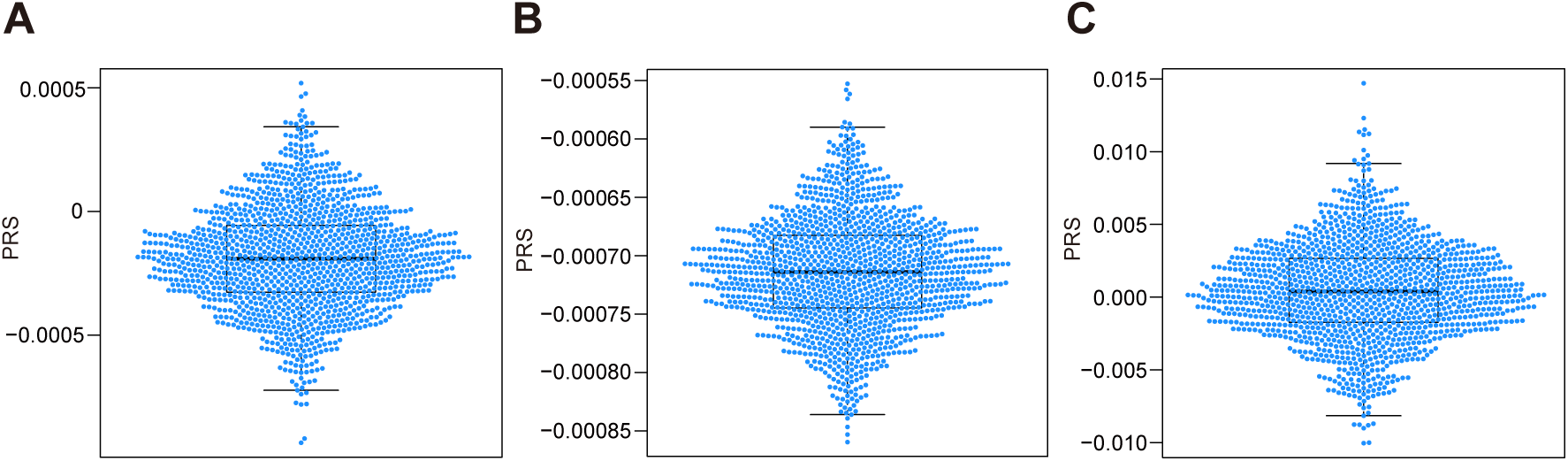
Distribution of constructed PRSs. Distribution of PRSs based on Jans2019 (**A**), Wigh2021 (**B**), and Shig2021 (**C**).

**Supplementary Figure S3.**
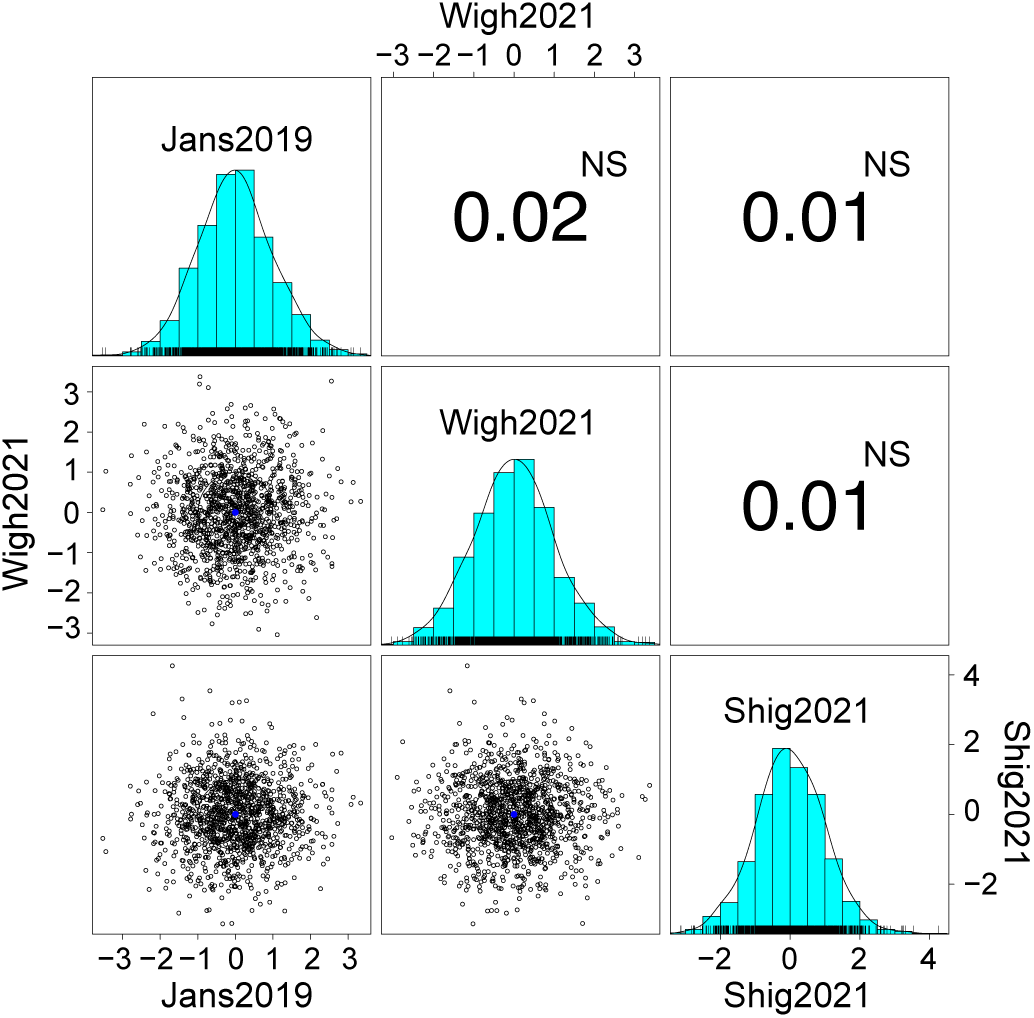
Correlation and distributions of three standardized PRSs. Correlation plot of three standardized PRSs. Histograms on the diagonal show the distribution of each PRS. Off-diagonal panels display Pearson’s correlation coefficients. Both axes represent standardized PRS values. The center of each distribution is marked by a blue point. NS indicates non-significant correlations.

## Supplementary Methods

### Brain imaging analysis in the Arao cohort

Brain MRI data were obtained in accordance with a previously reported protocol (Yoshiura et al., 2022). Imaging was conducted at two sites using 1.5-T scanners: Arao Municipal Hospital (Philips Ingenia CX Dual, Philips Healthcare, Amsterdam, Netherlands) and Omuta Tenryo Hospital (Signa HDxt Ver.23, GE Healthcare, Chicago, IL, USA). To account for potential inter-scanner variability, scanner type was included as a covariate in regression analyses, and no significant effects were detected. At Arao Municipal Hospital, MRI acquisition included high-resolution three-dimensional (3D) T1-weighted imaging (TR = 8.6 ms, TE = 4.0 ms, flip angle = 9°, matrix = 192 × 192, slice thickness = 1.2 mm), along with 3D T2-weighted, fluid-attenuated inversion recovery (FLAIR), and susceptibility-weighted imaging (SWI) sequences. At Omuta Tenryo Hospital, a comparable protocol was applied using 3D T1-weighted imaging (TR = 8.3 ms, TE = 3.4 ms, flip angle = 8°, matrix = 192 × 192, slice thickness = 1.2 mm), together with 3D T2-weighted, FLAIR, and T2*-weighted angiographic sequences. Volumetric analyses were performed using FreeSurfer version 7.0, following previously described procedures (Shima et al., 2024). Left and right hippocampal volumes were derived from 3D T1-weighted images.

